# Brain flexibility increases during the peri-ovulatory phase of the menstrual cycle

**DOI:** 10.1101/2023.06.19.23291595

**Authors:** Marianna Liparoti, Lorenzo Cipriano, Emahnuel Troisi Lopez, Arianna Polverino, Roberta Minino, Laura Sarno, Giuseppe Sorrentino, Fabio Lucidi, Pierpaolo Sorrentino

**Author notes:** These authors contributed equally (Marianna Liparoti, Lorenzo Cipriano).

## Abstract

The brain operates in a flexible dynamic regime, generating complex patterns of activity (i.e neuronal avalanches). This study aimed to describe how brain dynamics change according to menstrual cycle (MC) phases.

Brain activation patterns were estimated from resting state magnetoencephalography (MEG) scans, acquired women at early follicular (T1), peri-ovulatory (T2) and mid-luteal (T3) phases of MC. We investigated the functional repertoire (number of ways in which large bursts of activity spread through the brain) and the region-specific influence on large-scale dynamics across MC. Finally, we assessed the relationship between sex hormones and changes in brain dynamics.

A significantly larger number of visited configuration patterns, in T2 than in T1, in the beta frequency band was observed. No relationship between changes in brain dynamics and sex hormones was showed. Finally, we showed that, in the beta band, the left posterior cingulate gyrus and the right insula were more present in the functional repertoire in T2 than in T1, while the right pallidum was more present in T1 than in T2.

In summary, we showed a hormone independent increase of brain dynamics during the ovulatory phase. Moreover, we demonstrated that several specific brain regions play a key role in determining this change.

## Introduction

Human brain architecture enables efficient information processing, supporting complex cognitive and behavioural functions ^1^. Such complexity is underpinned by locally and globally integrated hierarchical and modular organisations ^2^, with simple components operating in different, nested spatio-temporal scales.

Network theory is a powerful approach to explain the functioning of a complex and integrated system such as the human brain ^3^. Accordingly, the brain has been conceptualised as a graph whose nodes are anatomical regions, and the edges are the connections (either structural or functional) among them. Exploiting this theory, structural and functional connectivity studies have attempted to measure anatomical connections (structural connectivity) and undirected statistical dependencies (functional connectivity) between pairs of brain regions ^1^, both in physiological and pathological conditions. However, most of the functional metrics used to this date assume stationarity of the brain, which is a convenient, yet inaccurate simplification ^4, 5^. In fact, brain activities are non-stationary, and generate bursts ^6^. This evidence highlights the importance of studying brain activity with different approaches that take into account dynamic variations in brain activity.

Studies on emergent properties of dynamic systems ^7, 8^ might suggest that the brain operates in a critical regime which optimises the information transmission, and supports the generation of complex spatio-temporal patterns. Large-scale brain activity is characterised by aperiodic bursts of activity, called *neuronal avalanches*. The neuronal avalanches generated in the human brain are typically not stereotyped and, rather, they reconfigure themselves in space and time, showing mutable behaviour ^8–11^. In this manuscript, we defined a neuronal avalanche operationally, as an event that begins when the activity of at least one region deviates from its baseline activity and ends when all regions restore their typical level of activity. An *avalanche pattern* is the set of all the brain areas that were recruited during the avalanche and the *functional repertoire* is the set of the unique patterns, which is interpreted as a measure of brain flexibility and, hence, of optimal brain functioning ^12^.

The menstrual cycle (MC) is a cyclic process originating from the coordinated variation of different sex hormones ^13, 14^, aimed at the reproductive process. However, the MC affects the body well beyond the reproductive organs, also inducing changes in the functioning of the central nervous system and in the women’s emotional and behavioural states. For this reason, researchers focused on neuroadaptive mechanisms that modulate brain structure and function during the MC. They demonstrated structural changes in the hippocampus, amygdala as well as in temporal and parietal regions ^15–17^. Furthermore, changes have been demonstrated also in brain connectivity, in particular in the brain topology which is related to phase of the MC and the corresponding hormonal changes ^18–22^ associated with clinically relevant conditions, such as the premenstrual syndrome or the premenstrual dysphoric disorder ^23, 24^. Although this large body of literature supports brain structural and functional change associated with the MC, to our knowledge few studies have focused on the possible changes of brain dynamics along the MC. In particular, De Filippi et al. ^25^, through the turbulence framework, demonstrated changes in whole-brain dynamics linked to ovarian hormones and menstrual cycle phases. Furthermore, Mueller et al. ^26^, through dynamic community detection techniques, demonstrated that hormonal changes during the menstrual cycle result in temporary and localised patterns of brain network reorganisation. However, these two studies are based on a densely sampled data set from a single participant. In this manuscript we use a larger sample of subjects, although with a lower sampling rate over time (ie. three time points, as explained), to clarify the role of the menstrual cycle on brain dynamics.

In the present study, we hypothesised that the brain dynamics cyclically changes along the MC. To verify our hypothesis, we evaluated the functional repertoire at three time points of MC (early follicular, peri-ovulatory and mid-luteal phases). Moreover, our study was aimed at verifying the hypothesis that possible modification in brain dynamics may be linked to changes in blood levels of sex hormones.

To test our hypothesis, we used a magnetoencephalography (MEG) system to measure the brain dynamic patterns of activation in resting state, in healthy naturally-cycling women without premenstrual symptoms and with no signs of anxiety and/or depression. In particular, we applied the *’’neuronal avalanches’’* framework to describe the variability of brain functional repertoire along the MC. Furthermore, during the three phases of the MC, blood samples were collected to determine hormone levels of estradiol, progesterone, luteinizing hormone (LH) and follicular-stimulating hormone (FSH) in order to test the hypothesis of a correlation between changes of brain dynamics and sexual hormones. Finally, we analysed the occurrence of brain regions in specific and shared functional repertoires.

## Methods

### Participants

For our study twenty-seven females were recruited (age and education 26.6 ± 5.1 and 17.3 ± 2.7 years, respectively). They were native Italian speakers, right-handed and heterosexual with a regular MC. At enrolment, all women signed informed consent. All the procedures were conducted in accordance with the Declaration of Helsinki, IV edition. The study was approved by the Local Ethics Committee of University of Naples “Federico II” (protocol n. 223/20). We selected women with regular MC (mean cycle length 28.4 ± 1.3 days), who did not use drugs or medicines, including the hormonal contraceptives (or other hormone regulating medicaments) during the last six month before the start of the study, since these drugs might affect both the central nervous system and the MC. We excluded participants who became pregnant in the last year, with a history of neuropsychiatric diseases or premenstrual dysphoric/depressive symptoms. Furthermore, the participants did not consume tobacco, alcohol and coffee in the 48h preceding the MEG recordings. To control for the influence of circadian rhythm, the time of testing varied no more than two hours between testing sessions. The Beck Depression Inventory (BDI)^27^ and Beck Anxiety Inventory (BAI)^28^, were used to test the mood and/or anxiety symptoms. Based on their cut-off (10 and 21 respectively), two females were excluded from the study, because the BDI test score had dropped below the cut-off.

### Experimental protocol

The women were acquired in three different time points of MC, at early follicular (cycle day 1-4, low estradiol and progesterone, T1), during the peri-ovulatory (cycle day 13-15, high estradiol, T2) and in mid-luteal (cycle day 21-23, high estradiol and progesterone, T3) phases. We applied the back-counting method to define the individual time points cycle. Self-reported onset of menses was used as a starting point and to estimate in an indirect way the peri-ovulatory and mid-luteal windows. At each time point of the cycle we performed the MEG recordings and the blood sample collection for the analysis of the hormones that act mainly during the menstrual cycle, which includes estradiol, progesterone, FSH and LH. During the early follicular phase, we also carried out a transvaginal pelvic ultrasonography examination. Finally, after the last MEG recording, a structural magnetic resonance imaging (MRI) was performed. To control for a possible session effect, we randomised the MC phase from which the three recording sessions started. For a full explanation of the procedures for analysis of hormone levels, pelvic ultrasound and MRI, see supplementary information (SI).

### Brain dynamics analysis

Brain magnetic signals were recorded through a MEG system, equipped with 163 magnetometers and placed in a shielded room ^29^. Before registration, the head position was identified, digitising the coordinates of four anatomical landmarks and four position coils ^30^. Each participant underwent two eyes-closed recordings of 3:30 min each. Electrocardiogram and electro-oculogram signals were acquired, to identify physiological artefacts ^31^. Brain signals were then cleaned through automated processes as reported in our previous article ^32^. The FieldTrip software tool ^33^, based on Mathworks® MATLAB, was used to implement principal component analysis (PCA), to reduce the environmental noise. To remove physiological artefacts such as cardiac noise or eye blinking (if present) independent component analysis (ICA) was used. For each participant, source reconstruction was performed through a beamforming procedure using the Fieldtrip toolbox, similarly to Pesoli et al. ^34^. In short, based on the native MRI, the volume conduction model proposed by Nolte^35^ was applied, and the Linearity Constrained Minimum Variance ^36^ beamformer was implemented to reconstruct time series related to the centroids of 116 regions-of-interest (ROIs), derived from the Automated Anatomical Labelling (AAL) atlas. We only considered the first 90 ROIs, excluding those corresponding to cerebellum, given that the reconstructed signal might be less reliable ^37^. The source-reconstructed signals were filtered in the classic frequency bands: delta (0.5–4.0 Hz), theta (4.0–8.0 Hz), alpha (8.0–13.0 Hz), beta (13.0–30.0 Hz), and gamma (30.0–48.0 Hz) ^38^.

We started from the time series of 90 ROIs to estimate the *Neuronal Avalanches* ^8^, defined as an event during which a large fluctuation of activity occurs above a certain established threshold value. This event begins when at least one region becomes active (i.e., above threshold) and continues as long as any region remains above the threshold and can involve both adjacent areas and areas of different hemispheres. The combination of all the brain regions that were involved in a specific avalanche is defined as “avalanche configuration” (or avalanche pattern). Therefore, before estimating the fluctuation of neuronal activity, we standardised each signal by calculating its z-score and establishing a threshold value equal to three standard deviations (z = ±3) ^39^. Hence, a region was considered as “active” only when its score was beyond the threshold. However, to demonstrate that the results are not strictly dependent on the chosen threshold, we repeated the analyses by setting two other cut-offs equal to + 10% and -10% of the initial threshold value (z = 3.3 and z = 2.7).

Dynamic analysis of neuronal signals requires the time series to be binned, to ensure we are observing a critical phenomenon. To select a suitable bin length, we computed the branching ratio σ ^40, 41^. In other words, for each time bin duration, for each subject, for each avalanche, the (geometrically) averaged ratio of the number of events (activations) between the subsequent time bin and that in the current time bin was calculated with the following formula:

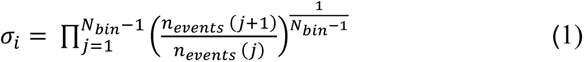

where σi is the branching parameter of the i-th avalanche in the dataset, Nbin is the total number of bins in the i-th avalanche, n events j is the total number of events in the j-th bin. Subsequently we (geometrically) averaged the results over all avalanches:

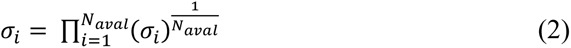

where N*aval* is the total number of avalanches in each participant’s dataset. In critical processes, a branching ratio ∼1 indicates critical processes with activity that is highly variable and nearly sustained, σ < 1 indicates subcritical processes in which the activity quickly dies out, and σ > 1 indicates supercritical processes in which the activity increases. While the branching ratio was close to one for all the bins that we tested, and the statistical results are confirmed for all bins, we report in the manuscript the results for bin = 4, since this was the closest to 1.

For the dynamic evaluation of neuronal activity, we estimated the following information: the *functional repertoire* composed by the number of unique avalanche configurations that was expressed during the recording; the *switches* between the state, represented by a crossing of the threshold in either direction between two consecutive time–bins; and the *regional influence on neuronal avalanches pattern*, which allows to role of each region within the MC phase-specific functional repertoires. Specifically, to estimate the regional influence on the neuronal avalanche patterns, the functional repertoire has been divided into two components: pattern that occurred in different phases of the MC, namely the “*shared repertoire*” and pattern that characterised the functional repertoire of one specific phase of the MC, defined as “*phase-specific repertoire*”. Subsequently, using the Kolmogorov-Smirnov test, we compared the distributions of the occurrences of the brain regions between shared and specific repertoires. Finally, we performed a permutation test to identify which brain regions occurred significantly more and in which repertoire (i.e., specific or shared). A brief description of the pipeline used and a schematic representation of neuronal avalanches are shown in Fig. 1.

**Figure 1.**
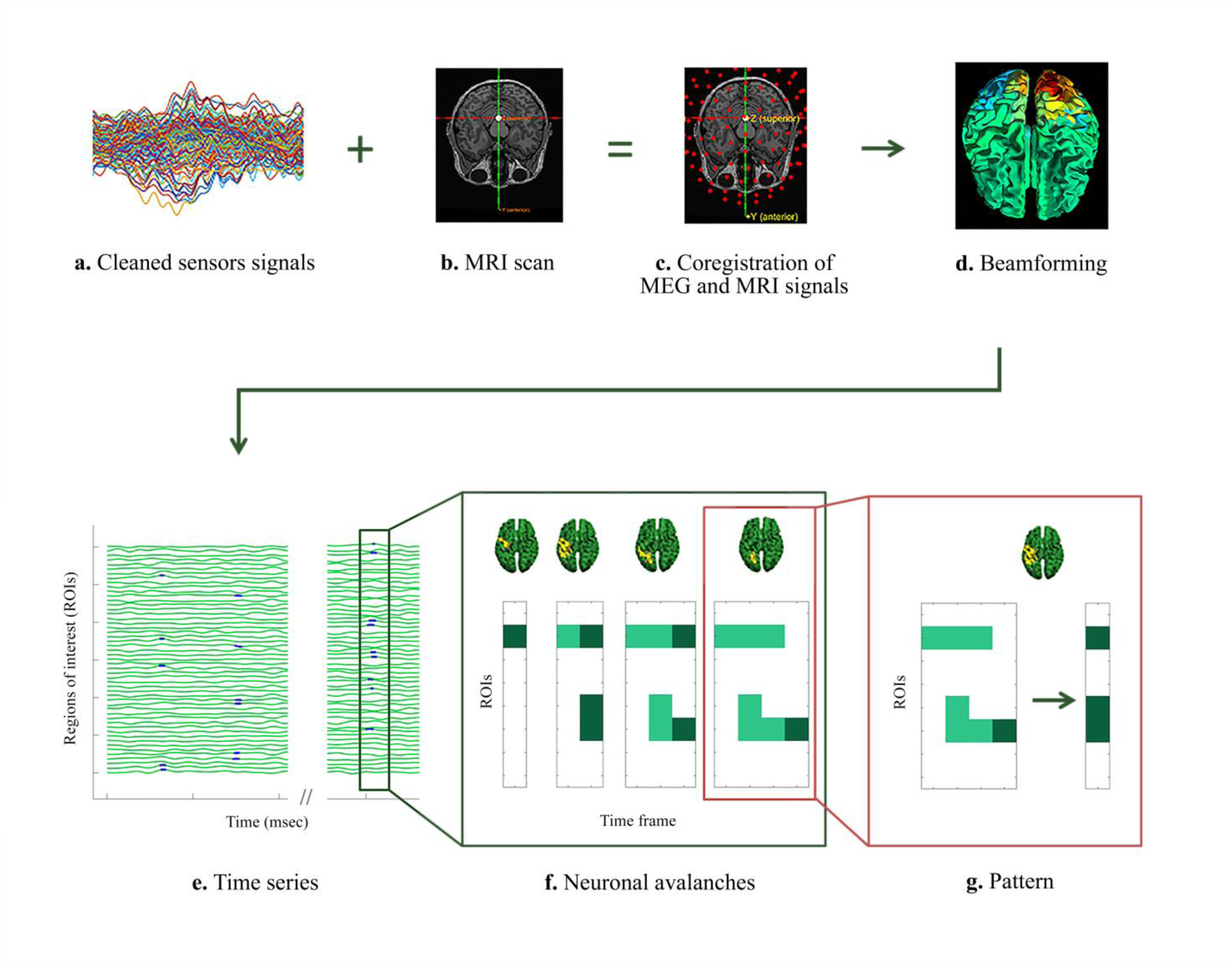
Data analysis pipeline and Neuronal Avalanches representation. (**a**) Magnetoencephalography (MEG) sensor-level signals after preprocessing and cleaning. (**b**) Structural magnetic resonance imaging (MRI). (**c**) Structural MRI, MEG sensors and the position of the participant’s head are co-registered in the same coordinate system. (**d**) Through a Beamformer algorithm, the time series of the sources are estimated in regions of interest within the brain according to a parcellation based on the ALL atlas. (**e**) In green, source-reconstructed time series of regions of interest. The dark green rectangle highlights the time interval in which a neuronal avalanche occurs. An avalanche is defined as an event that begins when at least one brain region exceeds the threshold value - equal to three standard deviations (z = ±3) away from its baseline activity - and ends when all regions have activity below the threshold. The blue dots included in the rectangle define the brain regions whose activity is above the threshold value in a given time interval (msec). (**f**) Frame by frame development of neuronal avalanches. Above, the set of all brain areas recruited during the avalanche (in yellow all the areas whose activity is above threshold and in green the areas whose activity is below threshold). At the bottom, the evolution in time of the avalanche is represented as follows: in each box, the dark green squares indicate the brain regions (ROIs) activated in a specific time frame, while the light green indicates the regions that were active in the previous frames. (**g**) Starting from the avalanche (to the left), we can calculate the patterns of activation (to the right), taking into consideration all the brain regions that were active during the avalanche, as represented in the brain plot above. The number of unique avalanche patterns defines the size of the functional repertoire and it is used as a proxy to estimate the flexibility of brain dynamics.

### Statistics

Statistical analysis was carried out using MATLAB (Mathworks®, version R2013a). We checked the normal distribution of variables using the Shapiro-Wilk test. To compare, in all frequency bands and all bins, the neuronal avalanches data among the three time points of the MC, we used the Friedman test. All the p values were corrected for multiple comparisons using the false discovery rate (FDR) ^42^. Subsequently, the post hoc analysis was carried out using the Wilcoxon signed rank test. The relationship between hormone blood levels and dynamic brain features was investigated through Spearman’s correlation test and multilinear regression analysis, using either the values of the specific phases, the values of the transition between two phases (Δ), or the overall variation of the hormones as assessed using the first component resulting from the PCA decomposition (see SI for more details). The statistical significance was defined as p < 0.05 after FDR correction.

## Results

### Brain dynamic analysis estimated by functional repertoire and switch rate

We measured the branching ratio of the time series binned with different lengths in order to observe the system in its critical state. For a threshold of z = 3, bin 4 showed a branching ratio σ = 1, hence our analyses were performed using time series in which each bin was obtained including four time points. However, the branching ratio remains very close to one for all the bins that were explored. Furthermore, the statistical results we obtained are stable across bins (bin t = 1, *p* = 0.023; bin t = 2, *p* = 0.026; bin t = 3, *p* = 0.026; bin t = 5, *p* = 0.026). The statistical comparison of the size of the functional repertoire across the three phases, based on the Friedman test, showed a significant difference among the functional repertoire of the three phases (χ2 (df = 2, N = 25) = 8.24, *p* = 0.016, *p*FDR = 0.048). In detail, the post hoc analysis showed a significantly higher number of visited configurations during the peri-ovulatory phase, as compared to the early follicular (*p* = 0.026) phase (Fig. 2). Furthermore, the results were confirmed even when testing different avalanche thresholds (*p* = 0.016 for z = 3.3, and *p* = 0.039 for z = 2.7).

**Figure 2.**
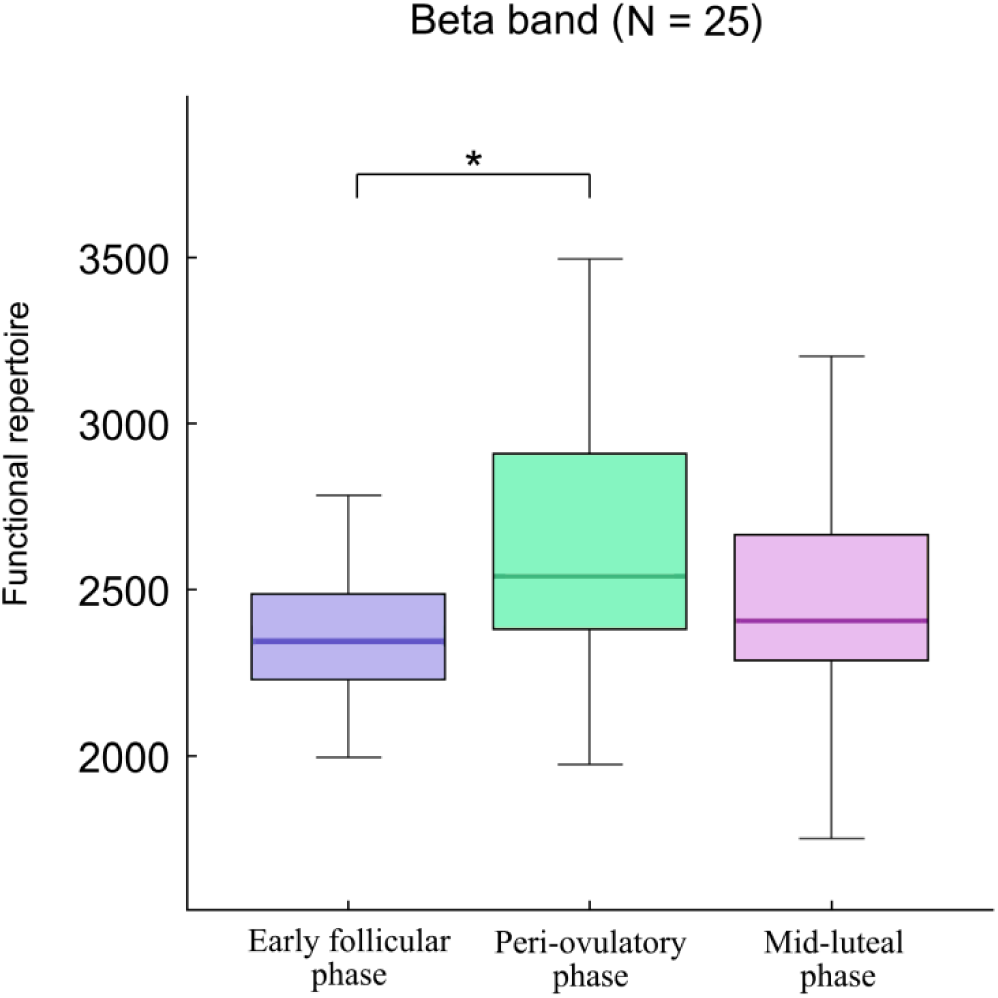
Neuronal Avalanches comparison. The box plots refer to the comparison of functional repertoires in the beta band, estimated in 25 women (N = 25) during the MC. In each box plot, the values are shown at early follicular, peri-ovulatory and mid-luteal phases. The upper and lower bound of the rectangles refer to the 25th to 75th percentiles, respectively. The median value is represented by a horizontal line inside each box, the whiskers extend to the 10th and 90th percentiles. The box plots show a significantly higher functional repertoire during the peri-ovulatory phase, as compared to the early follicular (p = 0.026) phase. Significance p value: *p < 0.05

With respect to the analyses of the switches, no statistically significant differences were found in any frequency band.

### Relationship between brain dynamics and blood sex hormones levels

Using multilinear modelling, Spearman correlations and principal component analyses, we investigated whether the changes in the size of the functional repertoire across the MC could be mediated by fluctuation of sex hormones we measured. All these analyses failed to demonstrate a relationship between variation in activation patterns and hormonal fluctuations. See SI for more details.

### Regional influence on neuronal avalanches pattern

With respect to the recruitment of specific brain regions in the phase-specific functional repertoire, we firstly compared the occurrence of specific regions on shared repertoire (i.e. the patterns that were present irrespective of the functional repertoire) with respect to the phase-specific repertoires. For this analysis, we focused on the phase-specific repertoires of those phases that demonstrated a statistically different number of patterns (i.e., early follicular and peri-ovulatory phases). The Kolmogorov-Smirnov test revealed a statistically significant difference in the brain regions occurrences distribution between shared and phase-specific repertoires of MC phases (p < 0.001) (Fig.3). Subsequently, we identified which brain regions occurred significantly more in the phase-specific repertoire than in the shared one, by performing a post-hoc analysis based on a permutation test. The results highlighted that, in the beta frequency band, the left posterior cingulate gyrus (PCG) (*p* = 0.047) and the right insula (*p* = 0.033) are recruited more often the functional repertoire of the peri-ovulatory phase, as compared to the one of early follicular phase. Furthermore, in the same frequency band, the right pallidum (*p* = 0.021) occurs more in the repertoire of the early follicular phase than in the one of the peri-ovulatory phase.

**Figure 3.**
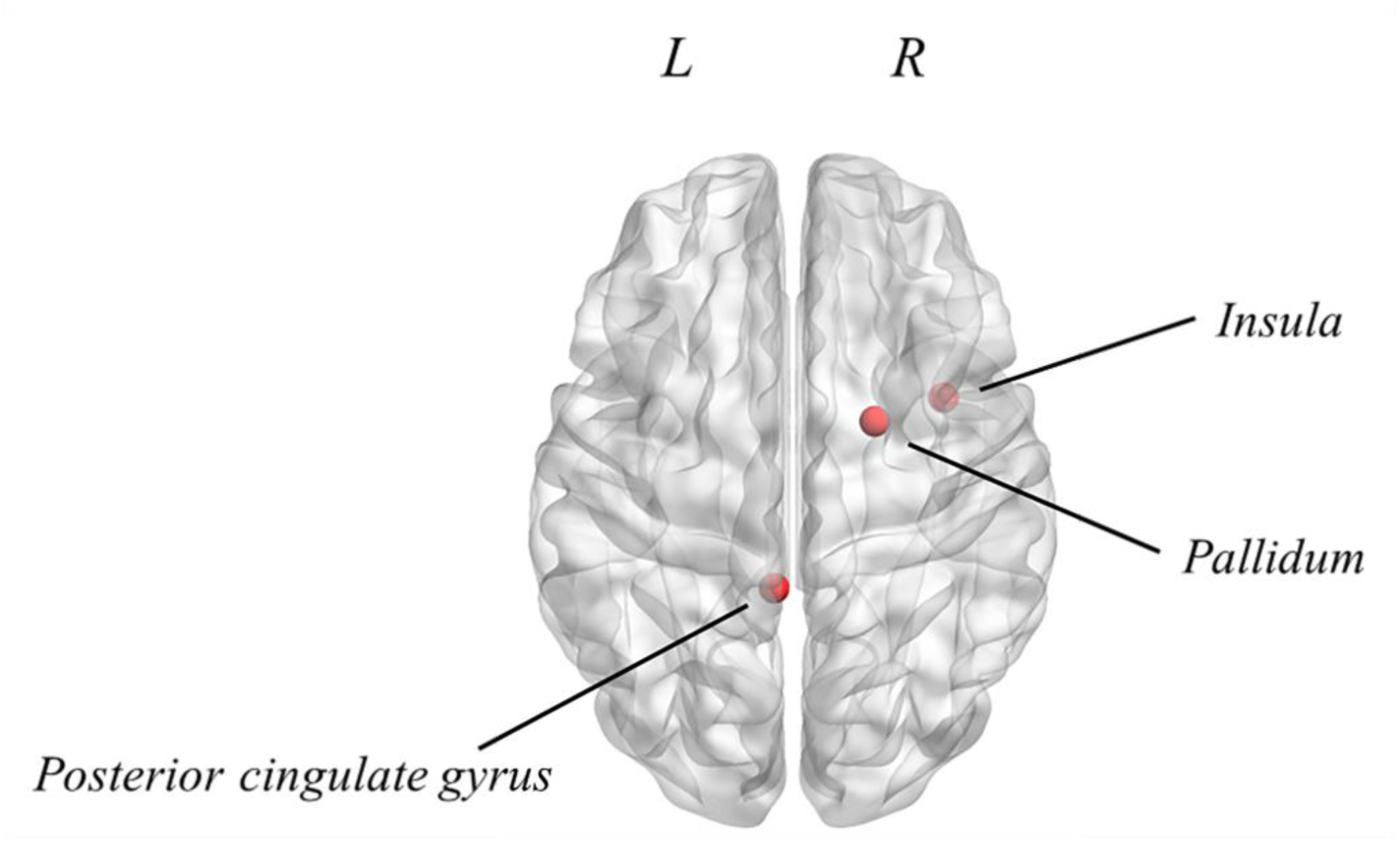
Mapping of brain regions occurring significantly more in the phase-specific unique avalanche patterns during the MC. In the beta frequency band, the left posterior cingulate gyrus (PCG) (*p* = 0.047) and the right insula (*p* = 0.033) occur more in the phase-specific repertoire of the peri-ovulatory phase than that of the early follicular phase. Furthermore, in the same frequency band, the right pallidum (*p* = 0.021) is recruited more often in the phase-specific repertoire of the early follicular phase, as compared to the one of the peri- ovulatory phase. The image was made using MatLab 2019a, including BraiNetViewer v. 1.62. Abbreviation: L (Left hemisphere); R (Right hemisphere).

## Discussion

In the present study, we set out to test the hypothesis that brain dynamics undergo significant variation across the MC, and we tested if those changes could be linked to physiological fluctuations of sex hormones that take place during the MC.

A robust literature suggests that the brain, in physiological conditions, changes its configuration across multiple temporal and spatial scales. The high number of configurations that are generated has been linked to the ability to adapt to a changing environment ^2, 3^. Here, we start from the evidence that the brain network topology, as observed in MEG, undergoes modifications across the MC, as demonstrated in our previous study ^43^. In the present work we aimed to investigate whether the MC affects the dynamical properties of the brain networks. We applied a recently implemented technique based on the physical concept of “*avalanche*” ^9^, through which we studied the variation of the *functional repertoire* during the MC as a proxy of the brain’s flexibility. While avalanches are typically related to the presence of a dynamics operating near a phase transition, our work does not aim at making a statement about the underlying dynamical regime. Rather, an operational definition of avalanches is adopted as a way to provide a read out of the flexibility of the dynamics.

Our results showed that the neuronal avalanches undergo profound rearrangements in different moments of the MC, as highlighted by the larger functional repertoire i observed during the peri-ovulatory phase as compared to the early follicular phase, in the beta frequency band. From a speculative perspective, during the fertility period, the woman’s brain may be required to reconfigure dynamically several patterns of activation to cope with several physiological changes which are related to adaptive behavioural modifications as well^44–46^.

It is interesting to note that the higher number of unique avalanches patterns occurred when the estradiol peaks. Our result is supported by the results by Muller et al. who observed a temporary dynamic reorganisation of the brain network during the ovulatory phase. Starting from these changes, we tested whether there we could demonstrate an association between sex hormones (in particular, estradiol, progesterone, FSH and LH) and the changes of functional repertoire that occur during the MC. To this aim, we firstly build multilinear models to predict the number of patterns of the functional repertoire in T1, in T2, as well as the observed changes between the two phases in Δ(T2-T1), from the hormonal levels. However, none of the models could significantly predict the functional repertoire (see Supplementary Fig. S1, Fig. S2 and Fig. S3). Subsequently, we explored the relationship between the number of unique avalanches and sex hormones, both individually and by extracting the first principal component of the hormone variations, using principal component analysis. The analysis was repeated in T1, T2 and Δ(T2-T1). As reported in SI, the analysis failed to demonstrate the association between sex hormones and the number of unique avalanches patterns. In summary, the analyses that we carried out did not demonstrate a linear relationship between the sex hormones assessed and the increase of functional repertoire observed during the peri ovulatory phase. Although there are evidences ^25, 26^ that demonstrate an association between changes in estradiol and progesterone concentrations and the whole-brain dynamic reorganisation on large scale, this was not replicated in the current study. It should be noted that the previous evidence is based on a dense sampling dataset obtained from a single participant. However, it must be considered that the regulation of the menstrual cycle depends on multiple different factors. For instance, the releasing of FSH and LH is determined by the gonadotropin-releasing hormone, secreted by the hypothalamus, whose regulation, in turn, depends on many factors determined by proteins, cytokines, and other physiological processes ^47^. Hence, the increase in the functional repertoire during the peri- ovulatory phase, expression of a fast large-scale process, may be the outcome of more complex mechanisms, involving broader physiological processes. Further analysis, considering a larger panel of molecular pathways, could clarify the topic.

We also analysed the number of switches that occurred, as a way to account for the possibility that the dynamics would not change qualitatively at the large-scale but, rather, the smaller functional repertoire would be derived from impaired local dynamics. However, we could not find any difference in terms of switch rates across the MC, demonstrating that the local dynamics is unchanged, despite the fact that the number of visited states (i.e. combinations of active regions) changes as a function of the phase of the MC. After observing the size of the functional repertoire, we investigated whether some brain regions recurred more than others in the functional repertoire. Therefore, we analysed the occurrences of the brain regions activations in shared and phase-specific patterns, and we observed that several brain regions were recruited more often in phase-specific functional repertoires. In particular, results showed an involvement of several areas of the limbic system including the PCG, the insula and the basal ganglia (pallidum). It is interesting to note that the limbic system is a complex system, whose relevance for controlling the emotional experience and in ensuring the survival and continuity of the species is well established ^48^.

In detail, the PCG might play a role in the regulation of brain dynamics during the peri- ovulatory phase. From a functional point of view, according to Leech and colleagues, the relevance of the PCG in the functional repertoire during the peri-ovulatory phase could depend on the role of this area in the assessment of the decision’s outcome ^49^.

Another area of the limbic system that seems to have greater relevance in the pattern of unique avalanches during the peri-ovulatory phase is the right insula. Although the insula is a small brain region, it is connected, at the large scale level, with several neuronal circuits involved in the regulation of visceral and somatic stimuli, in socio-emotional modulation and in cognitive processes ^50^. It was observed that its dysfunction, which might be due to a reduction in the volume of grey matter, has been associated with the subjective experience of pain during the menstrual cycle ^51^. One might speculate that the occurrence of this area in the functional repertoire of the peri-ovulatory phase could depend on the fact that the insula contributes to brain dynamics more prominently in the peri-ovulatory phase, perhaps favouring the expression of prosocial behaviours. However, this interpretation is purely speculative at this stage.

Finally, our study highlighted an occurrence of the basal ganglia (pallidum) in the functional repertoire along the menstrual cycle. Specifically, the right pallidum is recruited more often as part of the functional repertoire specific of the early follicular phase with respect to peri- ovulatory phase. Our result showed the role of the basal ganglia in shaping the dynamics and functional repertoire of the brain. The involvement of the basal ganglia during the menstrual cycle has been observed in previous studies, although these did not investigate brain dynamics. In particular, the basal ganglia appear involved in the inhibitory control processes, which is the ability to modulate (suppress or stimulate) cognitive processes in order to select one action over another ^52^. In this perspective, the activity of the basal ganglia seems to modulate the expression of impulsive behaviour.

In summary, this is the first MEG study to directly address dynamic activity of the brain across the MC. We showed that brain dynamics changes during the MC and, in particular, we observed increased functional repertoire during the peri-ovulatory phase. However, we could not find any correlation between the functional repertoire and the blood levels of the sex hormones. Further MEG studies measuring changes in brain dynamics are needed to confirm that the brain reorganises its activities across the MC as a function of physiological factors such as sex hormones. Finally, we investigated whether some brain regions recurred more than others in the functional repertoire of the time points of MC, and we found increased recruitment of areas belonging to the limbic system, whose role is crucial for the management of emotional and decision-making processes.

## Data Availability

The data that support the findings of this study are available from the corresponding author, PS, upon reasonable request.

## Declarations

### Author contributions

All authors had full access to all the data in the study and take responsibility for the integrity of the data and the accuracy of the data analysis. Conceptualization, M.L. and L.C.; Methodology, E.T.L. and P.S.; Investigation, M.L., L.C., E.T.L., P.S., L.S., R.M., and A.P.; Formal Analysis, M.L., L.C, E.T.L. and P.S.; Resources, G.S.; Writing – Original Draft, M.L., L.C.; Writing –Review & Editing, F.L.; Visualization, M.L., G.S. and P.S.; Project Administration, P.S.; Funding Acquisition, G.S

### Ethical statement

All procedures performed were in accordance with the ethical standards of the institutional research committee and with the ethical standards laid down in the 1964 Declaration of Helsinki and its later amendments.

### Informed consent and consent to participate

Written informed consent has been obtained from all participants.

