## Supplementary information for "Brain flexibility increases during the peri-ovulatory phase of the menstrual cycle"

**Marianna Liparoti[+1], Lorenzo Cipriano[+2], Emahnuel Troisi Lopez[3], Arianna Polverino[4], Roberta Minino[2], Laura Sarno[5], Giuseppe Sorrentino[2,3,4], Fabio Lucidi[1] and Pierpaolo Sorrentino[*3,6,7]**

[1] Department of Social and Developmental Psychology, Sapienza University of Rome, Rome, 00185, Italy

[2] Department of Motor Sciences and Wellness, University of Naples "Parthenope", Naples, 80133, Italy

[3] Institute for Diagnosis and Cure Hermitage Capodimonte, Naples, 80131, Italy

[4] Department of Neurosciences, Reproductive Science and Dentistry, University of Naples "Federico II", Naples, 80131, Italy

[5] Institut de Neurosciences des Systèmes, Aix-Marseille Université, Marseille, 13005, France

[6] Institute of Applied Sciences and Intelligent Systems, National Research Council, Pozzuoli, 80078, Italy

[7] Department of Biomedical Sciences, University of Sassari, Sassari, 07100, Italy

[+] These authors contributed equally

### Methods

*Ultrasonography examination*

The experimental protocol of the present study required that all enrolled women undergo ultrasound evaluation of the pelvis, in order to examine both the uterus and the ovaries and to detect the possible presence of abnormal findings such as endometrial polyps, myomas, ovarian cysts or other masses. Therefore, a transvaginal pelvic ultrasound was performed during the first follicular phase, using a 4-10 MHz (GE Healthcare, Milwaukee, WI). For evaluation, all women were in a lithotomy position with an empty bladder. Specifically, the uterus was scanned using the longitudinal and transverse planes and the thickness of the endometrium was measured at the widest point in the longitudinal plane. In addition, the number and diameter of the follicles were assessed for each ovary. None of the enrolled women had abnormal findings and endometrial thickness and follicle diameter were consistent with the menstrual phase.

*Magnetic Resonance Images (MRI) scan*

Twenty-five women underwent MRI scans. Recordings were performed by a 1.5-T Signa Explorer scanner equipped with an eight-channel parallel head coil (GE Healthcare, Milwaukee, WI, USA). Specifically, three-dimensional T1-weighted images (gradient-echo sequence Inversion Recovery prepared Fast Spoiled Gradient Recalled-echo, time repetition = 8.216 ms, TI = 450 ms, TE = 3.08 ms, flip angle = 12, voxel size = 1 × 1 × 1.2 mm3; matrix = 256 × 256) were acquired. Out of the 27 women recruited, 25 performed the MRI, while two participants refused it and a standard template was used for source reconstruction.

*Sex hormone assay*

During the early follicular, peri-ovulatory and mid luteal phases, all enrolled women underwent a venous blood sampling to assess the dosage of the following sex hormones: estradiol, progesterone, follicle stimulating hormone (FSH) and luteinizing hormone (LH). Specifically, all women were asked to observe 12 hours of fasting before the collection of blood samples. Whole blood samples were collected in S-Monovette tubes (Sarstedt), containing gel with coagulation activator to facilitate the separation of serum from the cellular fraction, according to the predetermined standard operating procedure [1]. For this purpose, samples were centrifuged at 4,000 rpm for 10 minutes, then serum was collected, aliquoted into 1.5 ml tubes (Sarstedt), and stored at -80°C until analysis. Hormonal assay was performed in order to assess whether women had a physiological range of the sex hormone levels. Specifically, we considered the following physiological hormone ranges: estradiol (range: 19.5-144.2 pg/ml (early follicular phase), 63.9-356.7 pg/ml (peri-ovulatory phase), 55.8-214. 2 pg/ml (mid-luteal phase), limit of detection: 11.8 pg/ml, coefficients of variation coefficients of variation on average: 1.9%, intra-assay coefficients of variation on average: 4.9%); progesterone (range: ND-1.4 ng/ml (early follicular phase), ND-2.5 ng/ml (peri-ovulatory phase), 2.5-28.03 ng/ml (mid-luteal phase), limit of detection: 0.2 ng/ml, inter-assay coefficients of variation average: 5.5%, intra-assay coefficients of variation average: 3.56%); LH (range: 1.9-12.5 mIU/ml [early follicular phase], 8.7-76.3 mIU/ml [peri-ovulatory phase], 0.5-16.9 mIU/ml [mid-luteal phase], limit of detection: 0.07 mIU/ml, coefficients of variation average: 2.3%; mean intra-test coefficients of variation: 2.5%) and FSH levels (range: 2.5-10.2 mIU/ml [early follicular phase], 3.4-33.4 mIU/ml [peri-ovulatory phase peri-ovulatory phase], 1.5-9.1 mIU/ml [mid-luteal phase], limit of detection: 0.3 mIU/ml, mean inter-assay coefficients of variation: 1.2%; mean intra-assay coefficients of variation: 1.9%). Hormone levels were measured by Advia Centaur XT Immunoassay System analyzer (Siemens) which uses competitive (estradiol) or direct (progesterone, FSH, LH) immunoassay and for quantification of reaction uses Chemiluminescent Acridinium Ester technology. The 2.5th and 97.5th percentiles were used to form reference limits with 90% confidence intervals, as provided by assay manufacturers [2]. The hormone blood levels at the three time points of the MC are reported in Table 1.

**Table 1.** Sex hormone blood levels

| **Sex hormone blood levels (N=27)** | | | | | | |
|---|---|---|---|---|---|---|
| **Hormones** | ***Early follicular (T1)*** | **Peri-ovulatory (T2)** | **Mid luteal (T3)** | $p_{FDR}$ **(T1 vs T2) †** | $p_{FDR}$ **(T2 vs T3) †** | $p_{FDR}$ **(T1 vs T3) †** |
| LH | 5.4 (± 2.3) | 16.4 (± 11.9) | 5.8 (± 4) | < 0.001 | < 0.001 | NS |
| FSH | 7.7 (± 1.7) | 8 (± 3.1) | 4.1 (± 1.4) | NS | < 0.001 | < 0.001 |
| Progesterone | 0.3 (± 0.1) | 1.1 (± 0.8) | 6 (± 3.4) | < 0.001 | < 0.001 | < 0.001 |
| Estradiol | 36.7 (± 14.1) | 142.3 (± 78.4) | 96.8 (± 39.1) | < 0.001 | < 0.05 | < 0.001 |

*Note:* Analysis of variance (ANOVA, †) for each hormone (luteinizing hormone (LH, mlU/ml), follicular stimulant hormone (FSH, mlU/ml), progesterone (ng/ml) and estradiol (pg/ml)) estimated in 27 women ($N$ = 27) at the early follicular (T1), peri-ovulatory (T2) and mid-luteal (T3) phases of the menstrual cycle (MC). The post hoc analysis between MC time points (T1 vs. T2, T2 vs. T3, and T1 vs. T3) was performed using a paired sample $t$ test. Data are given as mean concentrations (± standard deviation). Significance $p$ value: < 0.05, < 0.01, < 0.001. Abbreviations: FDR, false discovery rate; NS, no significant.

### Results

#### *Multilinear regression analysis*

In the present study we hypothesised that the brain dynamically changes its patterns of activation according to the phase of the MC, and these modifications may be mediated by sex hormones. To test our hypothesis we built a multilinear model to predict changes in the size of the functional repertoire from sex hormones blood levels of each phase of the MC (early follicular (T1) and peri-ovulatory (T2)) Fig. S1 and Fig. S2. Hence, the model was built considering the variations in the size of the functional repertoire (in T1 and T2) as the dependent variable, while the variations of estradiol, progesterone, LH, and FSH (in T1 and T2) were used as predictors. Furthermore, age and education were added in the model as nuisance variables. A leave-one-out cross-validation (LOOCV) technique was used in order to check the generalisation capacity of the model and its reliability. Specifically, we built the multilinear model excluding each time a different subject from the model, and verifying the predictive power of the model toward the size of the functional patterns of activity of the excluded subject (in T1 and T2). As shown in Fig. S1 and Fig. S2 the model failed to predict the size value of patterns of activity both in T1 and T2.

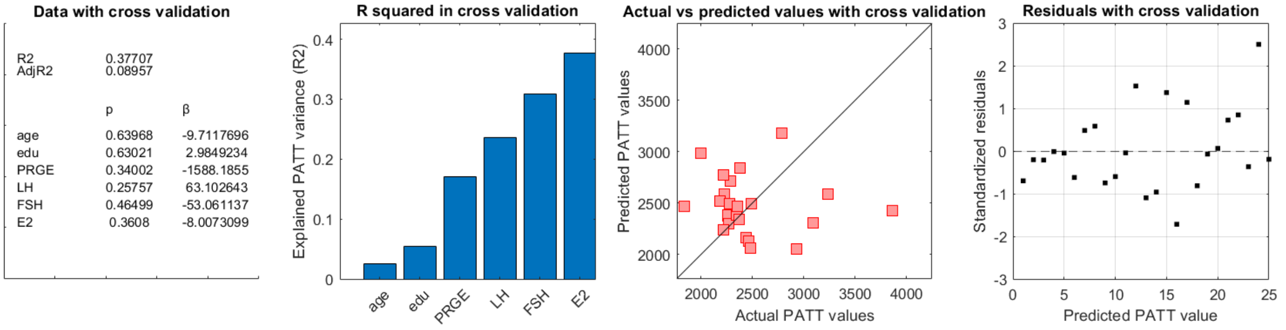


**Figure S1. Multilinear model with leave-one-out cross-validation (LOOCV) during early follicular phase (T1) of MC.** The model aims to predict the size variation of the pattern of the functional repertoire (PATT) starting from sex hormones blood levels at early follicular phase (T1) of the menstrual cycle (MC). (**a**) Data obtained with the cross validation; (**b**) Explained PATT variance (R2) of the model composed of two nuisance variables (age, education) and four predictors (progesterone, luteinizing hormone (LH), follicle-stimulating hormone (FSH), estradiol). (**c**) Scatter plot of the actual pattern of activity values versus the pattern of activity values predicted by the model with LOOCV. (**d**) Scatter plot of the standardised residuals (standardisation of the difference between actual and predicted (LOOCV) values). Significance p value: $*p < 0.05$, $**p < 0.01$, $***p < 0.001$; Abbreviation: β, Beta coefficient;

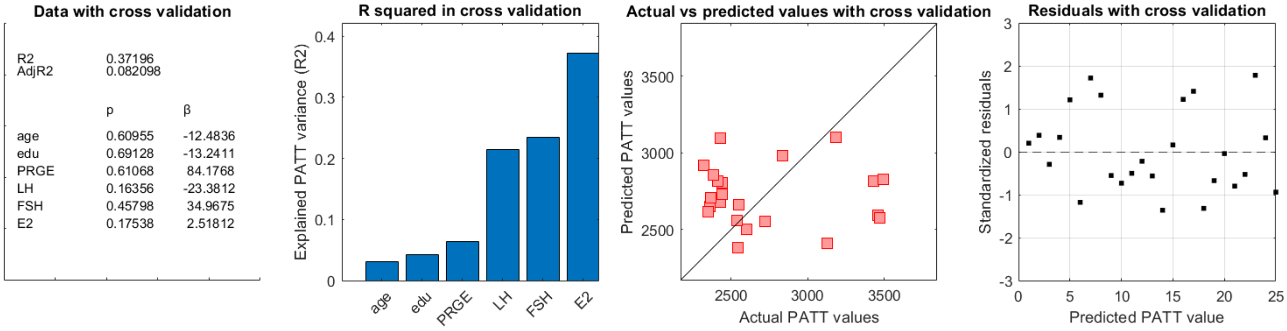


**Figure S2. Multilinear model with leave-one-out cross-validation (LOOCV) during peri-ovulatory phase (T2) of MC.** The model aims to predict the size variation of the pattern of the functional repertoire (PATT) starting from sex hormones blood levels at peri-ovulatory phase (T2) of the menstrual cycle (MC). (**a**) Data obtained with the cross validation; (**b**) Explained PATT variance (R2) of the model composed of two nuisance variables (age, education) and four predictors (progesterone, luteinizing hormone (LH), follicle-stimulating hormone (FSH), estradiol). (**c**) Scatter plot of the actual pattern of activity values versus the pattern of activity values predicted by the model with LOOCV. (**d**) Scatter plot of the standardised residuals (standardisation of the difference between actual and predicted (LOOCV) values). Significance p value: $*p < 0.05$, $**p < 0.01$, $***p < 0.001$; Abbreviation: β, Beta coefficient;

We also checked whether the model can  predict the changes in the size of the functional repertoire starting from the variation of the values of each sex hormone between two time points of MC Fig. S3. To this end we calculated the delta (Δ) values, expressed as the variations of the values of each parameter between two time points. Precisely, we considered the early follicular and peri-ovulatory time points of MC (Δ T2–T1, defined as the follicular phase) both for the brain dynamic parameters and the hormonal blood levels (estradiol, FSH, LH, and progesterone). As indicated in Fig. S3, the model failed to predict the size of the functional repertoire both in Δ T2–T1.

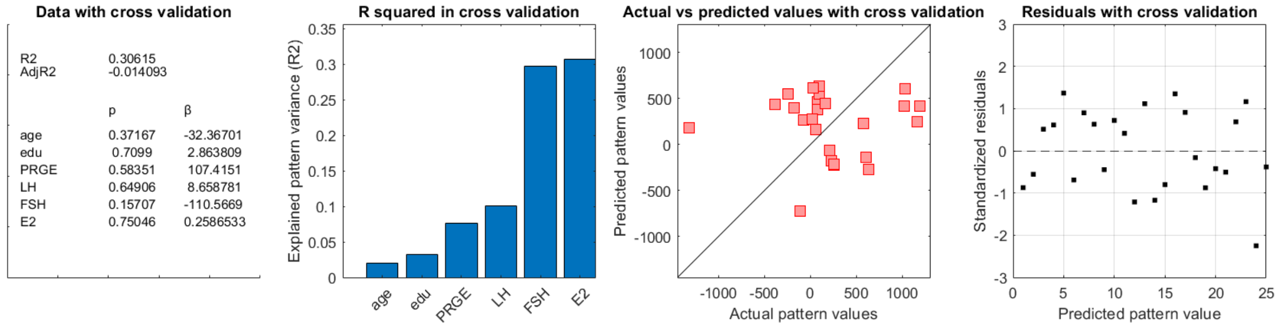


**Figure S3. Multilinear model with leave-one-out cross-validation (LOOCV) during the follicular phase (Δ T2–T1) of MC.** The model aims to predict the size variation of the pattern of the functional repertoire (PATT) starting from the variation (Δ) of sex hormones blood levels along early follicular and peri-ovulatory phase time points of the menstrual cycle (MC), calculated as (Δ T2–T1). (**a**) Data obtained with the cross validation; (**b**) Explained PATT variance (R2) of the model composed of two nuisance variables (age, education (edu)) and four predictors (progesterone, luteinizing hormone (LH), follicle-stimulating hormone (FSH), estradiol). (**c**) Scatter plot of the actual pattern of activity values versus the pattern of activity values predicted by the model with LOOCV. (**d**) Scatter plot of the standardised residuals (standardisation of the difference between actual and predicted (LOOCV) values). Significance p value: $*p < 0.05$, $**p < 0.01$, $***p < 0.001$; Abbreviation: β, Beta coefficient;

*Correlation analysis*

We performed a correlation analysis using the Spearman's correlation test, in order to investigate the relationship between dynamic brain changes and blood levels of the sex hormones. Specifically, we tested the relationship between the blood levels of the sex hormones and the dynamic brain changes during the time points of the MC (Fig. S4 and Fig. S5) in which the size of the functional repertoire varied  (early follicular (T1) and peri-ovulatory phases (T2)), and during the follicular phases, expressed as the variations of the values of each parameter between two time points of MC (Δ T2–T1) (Fig. S6). All Correlation analysis failed to demonstrate a linear relationship between dynamic brain changes and sex hormone blood levels.

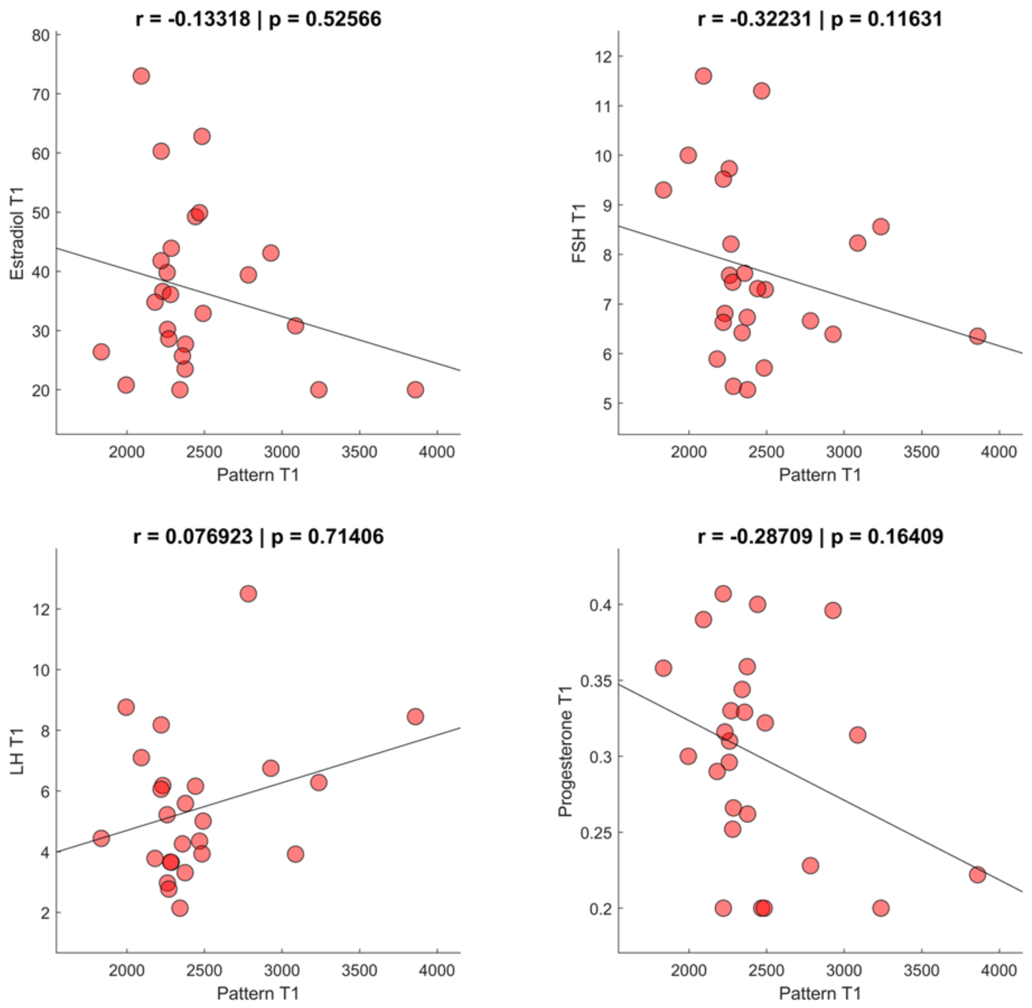


**Figure S4. Correlation between dynamic brain data and sex hormone blood levels during early follicular phase (T1) of MC.** Spearman's correlation between the size of the pattern of activity during the early follicular phase (Pattern T1) and sex hormones blood levels (Estradiol, Follicular Hormone Stimulation (FSH), Luteinizing Hormone (LH) and Progesterone) during the early follicular phase (T1) of the menstrual cycle. Significance p value: $*p < 0.05$, $**p < 0.01$, $***p < 0.001$;

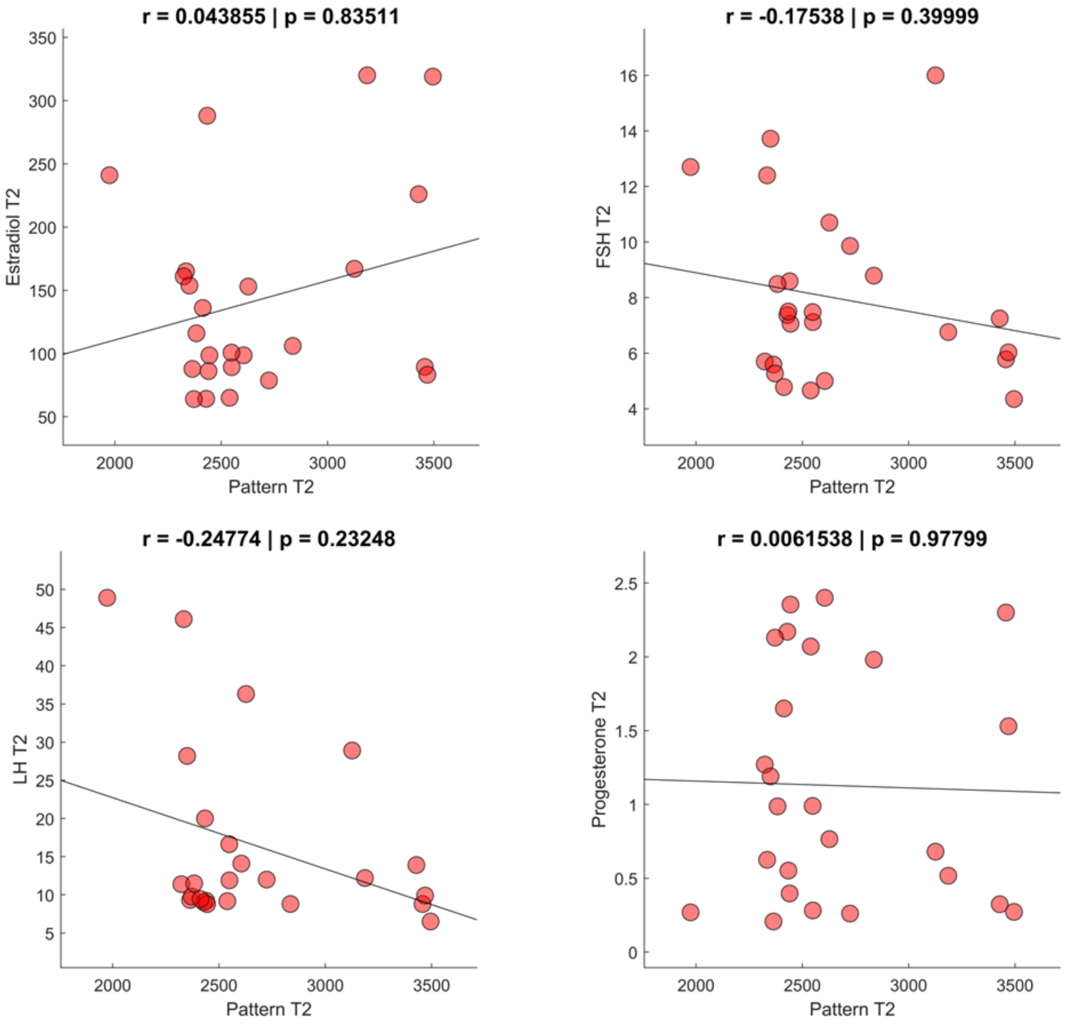


**Figure S5. Correlation between dynamic brain data and sex hormone blood levels during the peri-ovulatory phase (T2) of MC.** Spearman's correlation between the size of the pattern of activity during the peri-ovulatory phase (Pattern T2) and sex hormones blood levels (Estradiol, Follicular Hormone Stimulation (FSH), Luteinizing Hormone (LH) and Progesterone) during the peri-ovulatory phase (T2) of the menstrual cycle. Significance p value: *$p < 0.05$, **$p < 0.01$, ***$p < 0.001$;

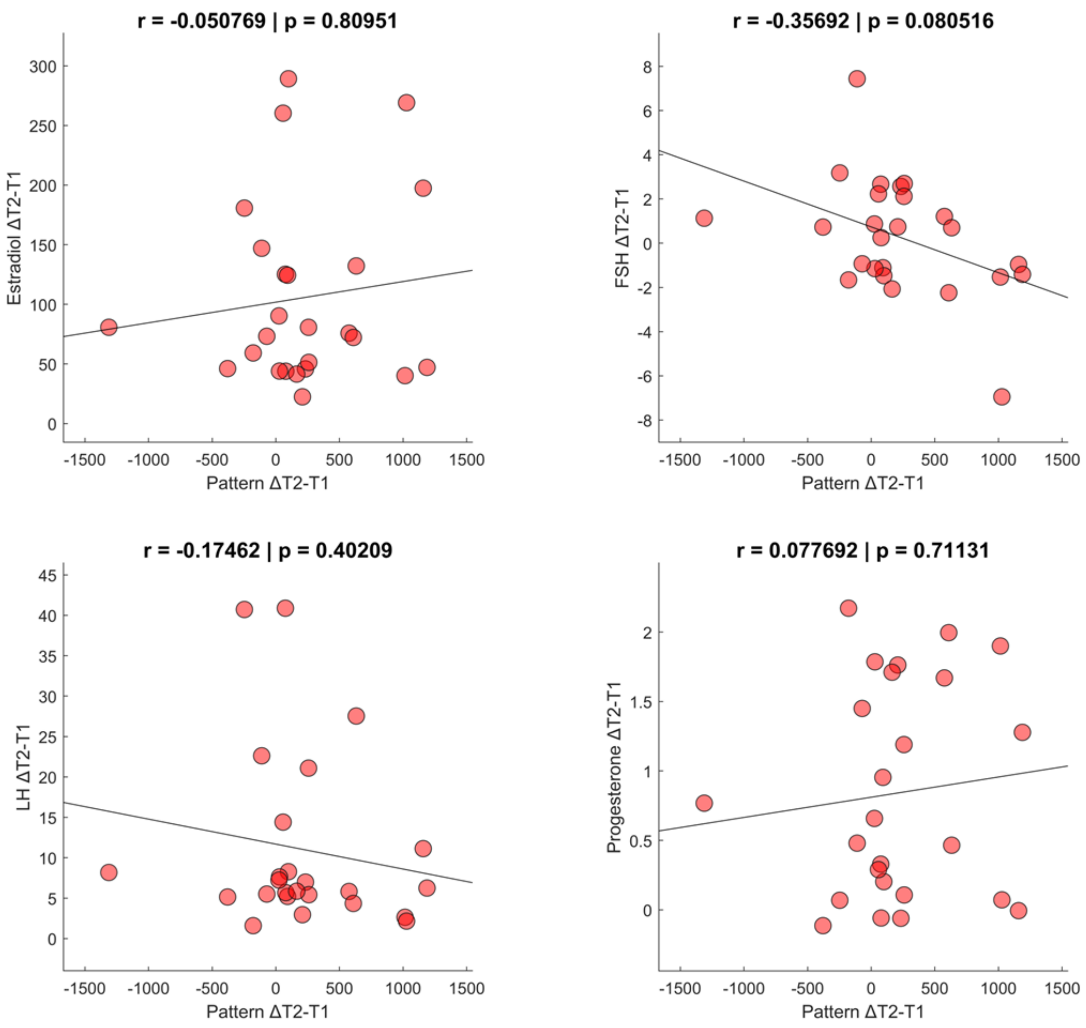


**Figure S6. Correlation between dynamic brain data and sex hormone blood levels during the follicular phase (Δ T2–T1) of menstrual cycle.** Spearman's correlation between the size of variation of the pattern of activity (Pattern Δ T2–T1) during two time points of menstrual cycle (MC) **(**the early follicular (T1) and peri-ovulatory (T2) phases of MC) and the variation of sex hormones blood levels (Estradiol, Follicular Hormone Stimulation (FSH), Luteinizing Hormone (LH) and Progesterone) during the same time points of MC. Significance p value: *$p < 0.05$, **$p < 0.01$, ***$p < 0.001$;

### *Principal component analysis*

Starting from the evidence that in our population the fluctuations of individual hormones in the blood levels do not correlate linearly with the variations in the functional repertoire, we supposed that the dynamic flexibility of neuronal avalanches might be influenced by a set of hormones that take part in the MC. For this purpose, we chose a multivariate statistical technique, the Principal Component Analysis (PCA), to reduce the dimensionality of our variables. Hence, we extracted the first principal component of our set of variables (i.e., E2, FSH, LH, PRGE) and, using the same methodology previously reported, we performed a correlation test between the projection over the first principal component and the size of the functional repertoire of both each time points of MC, in which a statistically significant difference was observed (early follicular (T1) and peri-ovulatory (T2)), and with the variation of each parameter between two points (Δ T2-T1, follicular phase). As shown in the Figure S7, principal component analysis also fails to show a relationship between the dynamic variations of the size of the functional repertoire observed during the MC and the hormones.

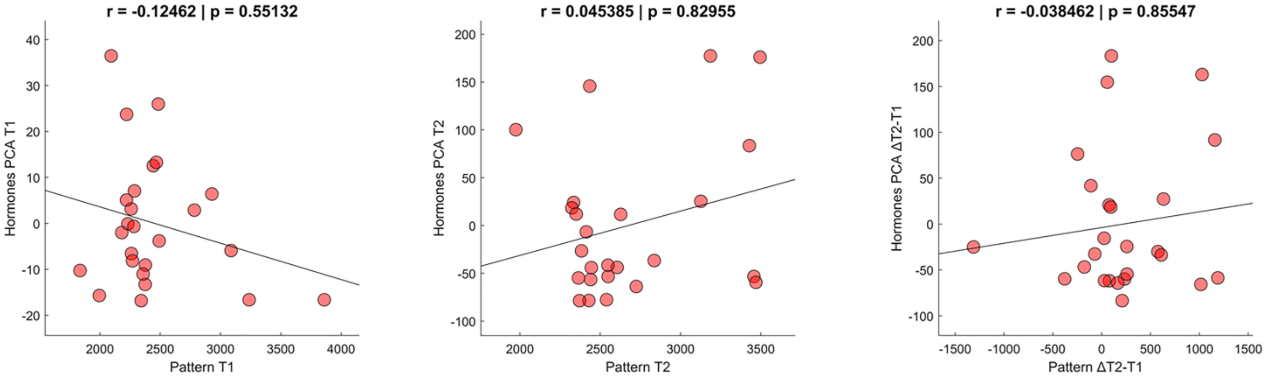


**Figure S7. Principal Component Analysis data. (a)** Correlation test between the size of the pattern of activity during the early-follicular phase (Pattern T1) and the first component of the principal component analysis (PCA) of sex hormones blood levels (i.e., E2 FSH LH PRGE) in the same time point of the MC. **(b)** Correlation test between the size of the pattern of activity during the peri-ovulatory phase (Pattern T2) and first component of the PCA of sex hormones blood levels in the same time point of the MC. **(c)** Correlation test between the variation of each parameter (patterns, and first component of the PCA of hormones blood levels) between two points (Δ T2-T1, follicular phase). Significance p value: *$p < 0.05$, **$p < 0.01$, ***$p < 0.001$;
